# Influenza vaccine effectiveness against influenza-associated hospitalizations in children, Hong Kong, November 2023 to June 2024

**DOI:** 10.1101/2024.08.30.24312831

**Authors:** So-Lun Lee, Mike Y. W. Kwan, Caitriona Murphy, Eunice L. Y. Chan, Joshua S. C. Wong, Sheena G. Sullivan, Malik Peiris, Benjamin J. Cowling

**Author notes:** Joint first authors with equal contribution (So-Lun Lee, Mike Y. W. Kwan, Caitriona Murphy).

## Abstract

We conducted a test negative study from November 2023 to June 2024, enrolling 4,367 children hospitalized with acute respiratory illness in Hong Kong. Among the children who tested negative for influenza virus and SARS-CoV-2, 56.8% had received influenza vaccination. Between November 2023 and March 2024, influenza A(H3N2) predominated and the VE against influenza A(H3N2) was estimated as 55% (95% CI: 29.6%, 71.8%). VE point estimates were higher for younger children than older children. In February to June 2024 influenza A(H1N1) predominated and VE against influenza A(H1N1) was 54% (95% CI: 33%, 69%) during this period. Influenza B circulated at low intensity throughout the 2023/24 season and VE against influenza B was 66% (95% CI: 42%, 80%). Since its introduction in 2018/19 the school-based influenza vaccination program has substantially increased vaccine uptake in children in Hong Kong and prevented influenza-associated hospitalizations.

## INTRODUCTION

Annual influenza vaccination is recommended for all individuals over 6 months of age. In Hong Kong, a school-based influenza vaccination program began in 2018 and progressively expanded to include children up to 18 years of age by 2022/23 [1]. The program for the 2023/24 season covered 70% of secondary schools, 95% of primary schools and 80% of kindergartens and childcare centers [2]. Children up to 6 years of age are also able to receive free or subsidized influenza vaccines via primary care physicians. Most vaccines administered in Hong Kong are inactivated split virion vaccines, and a small proportion of vaccinated children receive live attenuated influenza vaccines. The northern hemisphere vaccine formulation is used.

We have been monitoring influenza vaccine effectiveness (VE) against influenza-associated hospitalizations in children since 2009 [3,4]. Influenza was absent from Hong Kong between March 2020 and February 2023, when various public health and social measures were being used to control COVID-19 transmission [5]. Influenza circulation resumed in March 2023 and several influenza A epidemics have occurred since then [6,7], accompanied by steady circulation of influenza B. Here, we report estimates of influenza VE for the 2023/24 season including the second waves of A(H3N2) and A(H1N1) since the COVID-19 pandemic.

## METHODS

### Study design

We implemented a test-negative design study among children hospitalized with recent-onset acute respiratory illness at the Queen Mary Hospital on Hong Kong Island and the Princess Margaret Hospital in Kowloon. Based on their catchment populations, these two hospitals cover approximately 14% of all pediatric hospitalizations in Hong Kong. Eligible children were aged 6 months to 17 years with at least one respiratory symptom and a fever measuring ≥38°C. Information on influenza vaccination status was collected by interviewing parents or legal guardians using a standard questionnaire, and cross-checking responses with medical records. Relevant demographic and clinical information were obtained by chart review. At the time, all children admitted with respiratory symptoms were routinely tested on admission for influenza A and B virus, SARS-CoV-2 and other common respiratory viruses using an in-house multiplex PCR assay, and the FilmArray Respiratory Panel (BioFire/bioMérieux, Salt Lake City, UT).

Children were considered vaccinated if they had been fully vaccinated since 1 August 2023 and at least 2 weeks prior to hospitalization. Children were fully vaccinated if they were aged 9 years or older and had received one vaccine dose or, for children aged <9 years, if they had received two doses of influenza vaccine one month apart or one dose with a previous dose prior to August 2023 [8]. Because the second vaccine dose for infants is recommended at 7 months of age, we restricted enrolment to children aged at least 9 months of age. We reviewed influenza virus sequence data from GISAID to identify circulating virus clades during the study period. Our study protocol was approved by the Institutional Review Board of the Hospital Authority Hong Kong West Cluster and the Hospital Authority Kowloon West Cluster Research Ethics Committee. Verbal consent was obtained from the parents or legal guardians of participants.

### Statistical analysis

We determined relevant epidemic periods for influenza A subtypes by identifying contiguous periods during which there were at least 5 hospitalizations with that subtype each week. We used conditional logistic regression models to estimate conditional odds ratios for influenza positivity by vaccination status, adjusting for age (quadratic term), sex, prior year’s vaccination status and the presence of underlying medical conditions, conditioning by two-week periods of calendar time. VE was estimated as one minus the adjusted conditional odds ratio, multiplied by 100%. For each type/subtype analysis, VE estimates were made overall and stratified by age, and children were excluded from each control group if they tested negative for the type/subtype in question but positive for a different influenza type/subtype. Children testing negative for influenza but positive for SARS-CoV-2 were also excluded from each control group given the potential correlation between receipt of influenza vaccination and SARS-CoV-2 vaccination [9]. Statistical analyses were conducted in R version 4.2.2 (R Foundation for Statistical Computing, Vienna, Austria).

## RESULTS

During our study period, there was an epidemic of influenza A(H3N2) in November 2023 to early 2024 and then an influenza A(H1N1) epidemic in the spring of 2024. Influenza B circulated throughout the period with no distinct epidemic. Our focus in the following analyses is on estimation of influenza VE against hospitalizations associated with: (1) influenza A(H3N2) in the period November 2023 through March 2024; (2) influenza A(H1N1) in the period February 2024 through June 2024; (3) influenza B during the overall study period of November 2023 to June 2024 (Figure 1).

**Figure 1.**
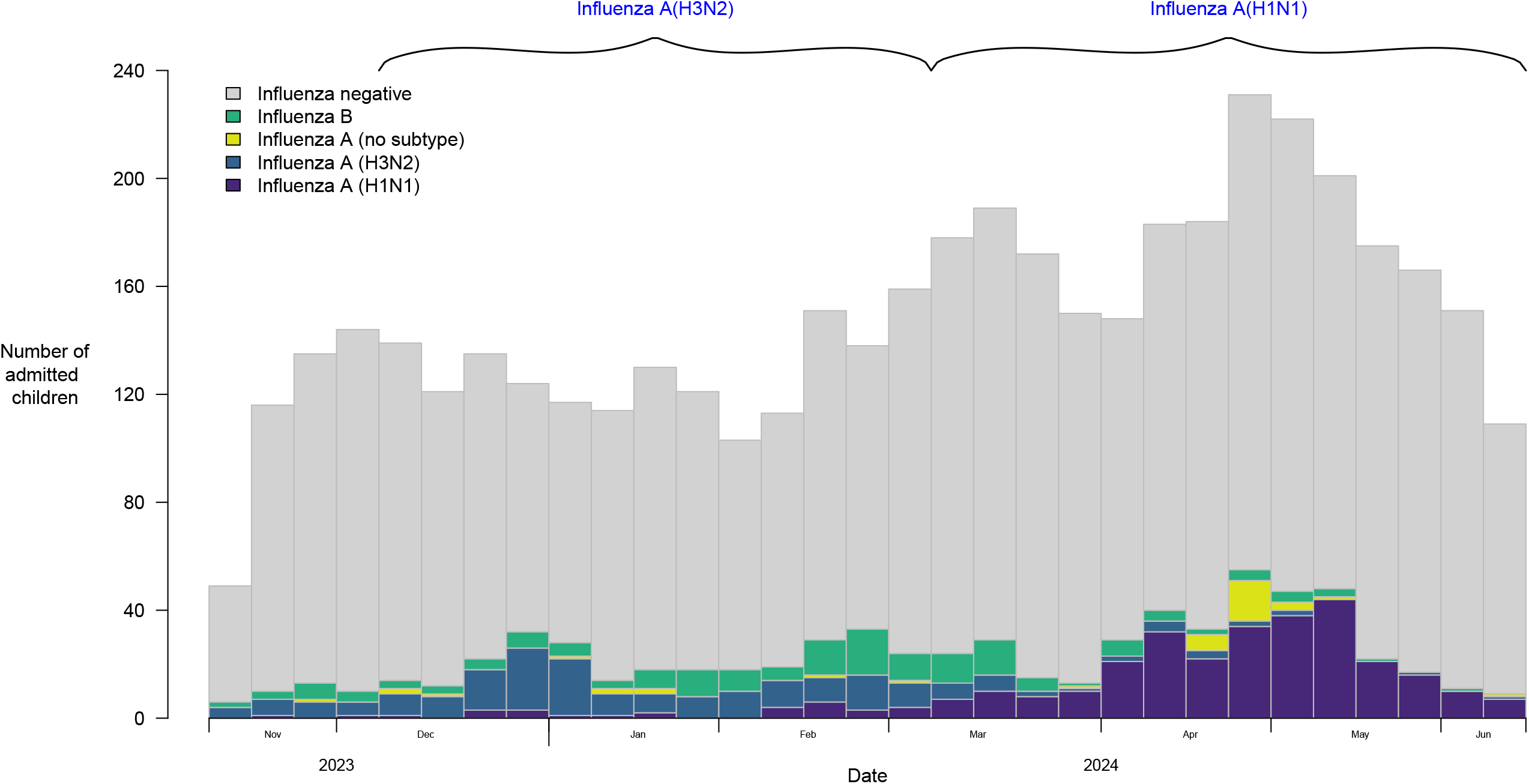
Number of children admitted to hospital and enrolled in this study from November 2023 to June 2024. The 2023-24 influenza A(H3N2) and influenza A(H1N1) epidemics were specified by the number of weekly hospitalizations and are indicated using brackets.

In total from 16 November 2023 to 12 June 2024, 4565 children were enrolled with acute respiratory illness. Among the influenza-negatives, there were 198 who tested positive for SARS-CoV-2 and were removed from analyses. Of the remaining 4367 children aged 9 months to 17 years, 709 (16%) tested positive for influenza. There were 2311 children who reported receipt of influenza vaccination (Table 1), including 2247 (97%) who received quadrivalent inactivated influenza vaccine and 51 (2%) that received quadrivalent live attenuated vaccine. Of the remaining 13 children, 8 received a trivalent vaccine and 5 received an unknown vaccination type.

**Table 1.** Characteristics of children hospitalized and enrolled in this study.

| Variable | Influenza positive<br>(n=709) | Influenza negative<br>and SARS-CoV-2<br>negative (n=3658) | p-value |
| --- | --- | --- | --- |
| **Male n (%)** | 410 (57.8) | 2015 (55.1) | 0.192 |
| **Age group n (%)** |  |  |  |
| 9 months to 3 years | 264 (37.2) | 1292 (35.3) | <0.001 |
| 4 to 8 years | 261 (36.8) | 1610 (44.0) |  |
| 9 to 17 years | 184 (26.0) | 756 (20.7) |  |
| **Underlying medical**<br>**conditions** |  |  |  |
| Lung diseases | 17 (2.4%) | 198 (5.4%) | 0.379 |
| Cardiac diseases | 3 (0.4%) | 18 (0.5%) |  |
| Other | 4 (0.6%) | 30 (0.8%) |  |
| **Receipt of influenza**<br>**vaccine n (%)** |  |  |  |
| 9 months to 3 years | 39 (14.8) | 466 (36.1) | 0.003 |
| 4 to 8 years | 117 (44.8) | 1119 (69.5) |  |
| 9 to 17 years | 78 (42.4) | 492 (65.1) |  |
| All ages | 234 (33.0) | 2077 (56.8) |  |

In the influenza A(H3N2) epidemic period, sequence data from GISAID indicated that a majority of circulating viruses fell in the 2a.3a.1 clade. These viruses are antigenically drifted from the 2023/24 northern hemisphere vaccine virus A/Darwin/9/2021(H3N2) and are more similar to the A/Thailand/8/2022, the vaccine virus recommended for the 2024 southern hemisphere vaccine. During these 17 weeks, 178 children tested positive for influenza A(H3N2), while 1941 tested negative for influenza A(H1N1) and B, and SARS-CoV-2. VE against influenza A(H3N2) was estimated to be 55% (95% confidence interval, CI: 30%, 72%) (Table 2). VE estimates were higher for younger children (Table 1).

**Table 2:** Influenza vaccine effectiveness estimates against hospitalization associated with influenza A(H3N2), A(H1N1) and B, overall and atified by age.

|  | Total | Influenza positive by PCR |  |  | Influenza negative and SARS-CoV-2 negative by PCR |  |  | Vaccine Effectiveness* |  |
| --- | --- | --- | --- | --- | --- | --- | --- | --- | --- |
|  |  | n vac | N | % | n vac | N | % | % | 95% CI |
| **Influenza A(H3N2)<sup>†</sup>** | 2119 | 70 | 178 | 39.3 | 1055 | 1941 | 54.4 | 55.4 | 29.6 to 71.8 |
| **Age stratified** |  |  |  |  |  |  |  |  |  |
| 9 months to 3 years | 794 | 10 | 59 | 16.9 | 250 | 735 | 34.0 | 60.3 | 13.0 to 81.9 |
| 4 to 8 years | 922 | 43 | 80 | 53.8 | 581 | 842 | 69.0 | 55.9 | 13.9 to 77.4 |
| 9 to 17 years | 403 | 17 | 39 | 43.6 | 224 | 364 | 61.5 | 27.0 | -149.5 to 78.7 |
| **Influenza A(H1N1)<sup>‡</sup>** | 2465 | 97 | 287 | 33.8 | 1308 | 2178 | 60.1 | 54.4 | 33.4 to 68.8 |
| **Age stratified** |  |  |  |  |  |  |  |  |  |
| 9 months to 3 years | 848 | 21 | 130 | 16.2 | 280 | 718 | 39.0 | 60.1 | 31.8 to 76.6 |
| 4 to 8 years | 1075 | 47 | 110 | 42.7 | 691 | 965 | 71.60 | 66.2 | 34.6 to 82.5 |
| 9 to 17 years | 542 | 29 | 47 | 61.7 | 337 | 495 | 68.1 | 26.2<sup>¶</sup> | -39.0 to 60.8 |
| **Influenza B<sup>§</sup>** | 3822 | 38 | 164 | 22.4 | 2077 | 3658 | 56.8 | 66.1 | 42.2 to 80.1 |
| **Age stratified** |  |  |  |  |  |  |  |  |  |
| 9 months to 3 years | 1343 | 3 | 51 | 5.9 | 466 | 1292 | 36.1 | 84.1 | 45.8 to 95.3 |
| 4 to 8 years | 1657 | 17 | 47 | 36.2 | 1119 | 1610 | 69.5 | 58.2 | -2.1 to 82.9 |
| 9 to 17 years | 822 | 18 | 66 | 27.3 | 492 | 756 | 65.1 | 44.7 | -62.4 to 81.2 |

\*Adjusted for age, sex, prior vaccination and underlying conditions
<sup>†</sup>Influenza A(H3N2) VE was estimated during the period 19 November 2023 to 23 March 2024
<sup>‡</sup>Influenza A(H1N1) VE was estimated during the period 25 February to 12 June 2024
<sup>§</sup>Influenza B VE was estimated during the period 16 November 2023 to 12 June 2024
<sup>¶</sup>Adjusted for age, sex and underlying conditions but not for prior vaccination status due to insufficient sample size

Sequence data for the influenza A(H1N1) epidemic from February to June 2024 indicated the predominant circulating clade was 5a.2a, similar to the A/Victoria/4897/2022 influenza A(H1N1) vaccine virus used in the 2023/24 northern hemisphere vaccine. During this period, 287 children tested positive for influenza A(H1N1). Estimated VE against influenza A(H1N1) was 54% (95% CI: 33%, 69%) overall, and higher in the younger children. Over the whole study period, 164 children were hospitalized with influenza B. The estimated VE against influenza B was 66% (95% CI: 42%, 80%) overall, and higher for children 9 months to 3 years (Table 1).

## DISCUSSION

We estimated VE against influenza A(H3N2) of 55% (95% CI: 30%, 72%) which was the same as the VE estimate of 55% (95% CI: 20%, 74%) for children 0-17 years reported by the US New Vaccine Surveillance Network (NVSN) for outpatients [10]. We are not aware of other published VE estimates against A(H3N2) specifically for children for the 2023/24 season. Our estimates are slightly higher than the interim all-age VE point estimates against influenza A(H3N2) of 30% (95% CI: -3%, 54%) in primary care and 14% (95% CI: -32%, 43%) in hospitals in Europe [11].

Our observed VE point estimate for A(H3N2) was also higher than in the preceding 2022/23 season in Hong Kong, which had the same vaccine formulation for influenza A(H3N2) and where the circulating influenza A(H3N2) also fell in the 2a.3a.1 clade [7]. Waning immunity was postulated to be a contributing factor to the low VE point estimate in 2022/23 against influenza A(H3N2) (VE: 14%; 95% CI: -29%, 43%), as that epidemic peaked more than six months after the usual vaccination period from October and November [7]. In contrast the A(H3N2) epidemic in 2023/24 occurred soon after vaccines had been administered.

The VE against influenza A(H1N1) in Hong Kong was estimated as 54% (95% CI: 33%, 69%), which was slightly lower than the estimate reported from the NVSN for inpatient children 6 months to 17 years (VE: 60%; 95% CI: 32%, 77%) [10] and from Canada for children 1-19 years (VE: 68; 95% CI: 42%, 83%) [12], and substantially lower than the VE reported among children in primary care in Europe (VE: 85%; 95% CI: 71%, 93%) [11]. Sequences uploaded to GISAID from the public health laboratory in Hong Kong indicated clade 5a.2a was circulating from March 2024. The WHO vaccine recommendation for influenza A(H1N1) changed clade from 5a.2a for the 2022/23 season to 5a.2a.1 for the 2023/24 season. Influenza A(H1N1) strains detected in Europe were more similar to the 2023/24 vaccine strain which may contribute to the higher VE estimated there.

Influenza B circulated at relatively low levels throughout the season. We estimated a VE against influenza B as 66% (42%, 80%). Estimates from the US NVSN outpatients reported a similar estimate for children of 64% (95% CI: 47%, 75%), but another three US outpatient networks reported higher VE against influenza B for children of 79% (95% CI: 71, 85; VISION), 89% (95% CI: 70, 97; US Flu VE) [10], and 79% (95% CI: 76%, 82%; California) [13]. Surveillance networks in Europe and Canada did not report a VE estimate against influenza B due to small sample sizes [11,12].

Hong Kong has experienced a longer period of influenza circulation in the 2023/24 season, lasting 28 weeks from mid-January [14] compared to 12 to 14 weeks for the same period in previous years [15], or compared to an average of 22 weeks for the 2013/14 to 2018/19 influenza seasons [16] that included circulation before January. Disruption to influenza seasonality after the COVID-19 pandemic has been reported elsewhere [17,18]. Whether or how quickly influenza patterns will return to pre COVID-19 seasonality remains to be seen.

Influenza vaccination coverage in Hong Kong has been higher in the 2023/24 season compared to previous seasons, with 49% of children under 6 years of age and 68% of children 6 to 12 years of age vaccinated this year [1]. This is an increase from 38% and 60% in the 2022/23 season for the same age groups. Similarly, among the controls in our study the vaccination coverage for children under 6 years and 6-12 years has increased from 8% and 12% respectively in 2009 to 2014 [3], to 14% and 15% in 2015/16 [19], to 30% and 59% in 2022/23 [6], and now to 43% and 70% in this study. In particular, influenza vaccine uptake in children has increased very substantially since the introduction of school-based vaccination in 2018/19 [1]. We estimated the direct effect of vaccination, but this school-based vaccination program may also have indirect benefits to unvaccinated and vaccinated children by reducing the overall prevalence of infection.

Some limitations of our study include a small sample size when stratifying estimates by age group resulting in low precision, particularly for the older age group which experienced fewer hospitalizations. Second, multiplex rapid antigen tests that detect several respiratory viruses including influenza are now readily available in Hong Kong [20] which could impact hospitalization rates if parents sought care or were prescribed antivirals based on timely rapid test results. Use of rapid tests might also be correlated with receipt of vaccination. In both scenarios, the use of rapid tests may alter healthcare seeking behavior and cause selection bias in test-negative studies.

In conclusion, during a prolonged influenza season in Hong Kong we estimated influenza VE against hospitalizations during a 17-week influenza A(H3N2) epidemic as 55% (95% CI: 30%, 72%) and during a 17-week influenza A(H1N1) epidemic as 54% (95% CI: 33%, 69%). Influenza B circulated throughout and the VE against influenza B was 66% (95% CI: 42%, 80%). School-based vaccination has led to increases in influenza vaccination coverage in children and reduced the burden of influenza-associated hospitalizations in children in Hong Kong.

## Data Availability

All data produced in the present study are available upon reasonable request to the authors

## ACKNOWLEDGMENTS

The authors thank colleagues at Queen Mary Hospital and Princess Margaret Hospital for facilitating participant enrolment. The authors thank Julie Au for technical support.

## REFERENCES

[1] Centre for Health Protection, Government of the Hong Kong SAR. Statistics on Vaccination Programmes in the Past 3 years 2024. https://www.chp.gov.hk/en/features/102226.html (accessed July 11, 2024).

[2] Wong WH-S, Peare S, Lam HY, Chow CB, Lau YL. The estimated age-group specific influenza vaccine coverage rates in Hong Kong and the impact of the school outreach vaccination program. Hum Vaccin Immunother 2022;18:1–5.

[3] Cowling BJ, Chan K-H, Feng S, Chan ELY, Lo JYC, Peiris JSM, et al. The effectiveness of influenza vaccination in preventing hospitalizations in children in Hong Kong, 2009-2013. Vaccine 2014;32:5278–84.

[4] Chua H, Kwan MYW, Chan ELY, Wong JSC, Peiris JSM, Cowling BJ, et al. Influenza vaccine effectiveness against influenza-associated hospitalization in children in Hong Kong, 2010-2020. Vaccine 2021;39:4842–8.

[5] Xiong W, Cowling BJ, Tsang TK. Influenza Resurgence after Relaxation of Public Health and Social Measures, Hong Kong, 2023. Emerg Infect Dis 2023;29:2556–9.

[6] Cowling BJ, Kwan MYW, Murphy C, Chan ELY, Wong JSC, Sullivan SG, et al. Influenza Vaccine Effectiveness Against Influenza-Associated Hospitalization in Hong Kong Children Aged 9 Months to 17 Years, March-June 2023. J Pediatric Infect Dis Soc 2023;12:586–9.

[7] Murphy C, Kwan MYW, Chan ELY, Wong JSC, Sullivan SG, Peiris M, et al. Influenza vaccine effectiveness against hospitalizations associated with influenza A(H3N2) in Hong Kong children aged 9 months to 17 years, June-November 2023. Vaccine 2024;42:1878–82.

[8] Grohskopf LA, Blanton LH, Ferdinands JM, Chung JR, Broder KR, Talbot HK, et al. Prevention and Control of Seasonal Influenza with Vaccines: Recommendations of the Advisory Committee on Immunization Practices - United States, 2022-23 Influenza Season. MMWR Recomm Rep 2022;71:1–28.

[9] Doll MK, Pettigrew SM, Ma J, Verma A. Effects of Confounding Bias in Coronavirus Disease 2019 (COVID-19) and Influenza Vaccine Effectiveness Test-Negative Designs Due to Correlated Influenza and COVID-19 Vaccination Behaviors. Clin Infect Dis 2022;75:e564–71.

[10] Frutos AM, Price AM, Harker E, Reeves EL, Ahmad HM, Murugan V, et al. Interim Estimates of 2023-24 Seasonal Influenza Vaccine Effectiveness - United States. MMWR Morb Mortal Wkly Rep 2024;73:168–74.

[11] Maurel M, Howard J, Kissling E, Pozo F, Pérez-Gimeno G, Buda S, et al. Interim 2023/24 influenza A vaccine effectiveness: VEBIS European primary care and hospital multicentre studies, September 2023 to January 2024. Euro Surveill 2024;29. 10.2807/1560-7917.ES.2024.29.8.2400089.

[12] Skowronski DM, Zhan Y, Kaweski SE, Sabaiduc S, Khalid A, Olsha R, et al. 2023/24 midseason influenza and Omicron XBB.1.5 vaccine effectiveness estimates from the Canadian Sentinel Practitioner Surveillance Network (SPSN). Euro Surveill 2024;29. 10.2807/1560-7917.ES.2024.29.7.2400076.

[13] Zhu S, Quint J, León TM, Sun M, Li NJ, Tenforde MW, et al. Interim Influenza Vaccine Effectiveness Against Laboratory-Confirmed Influenza - California, October 2023-January 2024. MMWR Morb Mortal Wkly Rep 2024;73:175–9.

[14] Centre for Health Protection, Government of the Hong Kong SAR. CHP announces end of influenza season 2024. https://www.info.gov.hk/gia/general/202407/25/P2024072500406.htm (accessed August 23, 2024).

[15] Centre for Health Protection, Government of the Hong Kong SAR. COVID-19 & Flu Express Volume 2, Number 28 (published on Jul 18, 2024) 2024. https://www.chp.gov.hk/files/pdf/covid_flux_week28_18_7_2024_eng.pdf (accessed August 23, 2024).

[16] Wong N-S, Leung C-C, Lee S-S. Abrupt Subsidence of Seasonal Influenza after COVID-19 Outbreak, Hong Kong, China. Emerg Infect Dis 2020;26:2753–5.

[17] Skowronski DM, Chuang ES, Sabaiduc S, Kaweski SE, Kim S, Dickinson JA, et al. Vaccine effectiveness estimates from an early-season influenza A(H3N2) epidemic, including unique genetic diversity with reassortment, Canada, 2022/23. Euro Surveill 2023;28. 10.2807/1560-7917.ES.2023.28.5.2300043.

[18] Wang MH, Hu ZX, Feng LZ, Yu HJ, Yang J. [Epidemic trends and prevention and control of seasonal influenza in China after the COVID-19 pandemic]. Zhonghua Yi Xue Za Zhi 2024;104:559–65.

[19] Cowling BJ, Kwan MYW, Wong JSC, Feng S, Leung C-W, Chan ELY, et al. Interim estimates of the effectiveness of influenza vaccination against influenza-associated hospitalization in children in Hong Kong, 2015-16. Influenza Other Respi Viruses 2017;11:61–5.

[20] Murphy C, Mak L, Cheng SMS, Liu GYZ, Chun AMC, Leung KKY, et al. Diagnostic performance of multiplex lateral flow tests in ambulatory patients with acute respiratory illness. Diagn Microbiol Infect Dis 2024;110:116421.

